# COVID-19 vaccine effectiveness against laboratory confirmed symptomatic SARS-CoV-2 infection, COVID-19 related hospitalizations and deaths, among individuals aged 65 years or more in Portugal: a cohort study based on data-linkage of national registries February-September 2021

**DOI:** 10.1101/2021.12.10.21267619

**Authors:** Ausenda Machado, Irina Kislaya, Ana Paula Rodrigues, Duarte Sequeira, João Lima, Camila Cruz, Pedro Pinto Leite, Carlos Matias-Dias, Baltazar Nunes

## Abstract

**Background:** Using data from electronic health registries, this study intended to estimate the COVID-19 vaccine effectiveness in the population aged 65 years and more, against symptomatic infection, COVID-19 related hospitalizations and deaths, overall and by time since complete vaccination.

**Methods:** We stablished a cohort of individuals aged 65 and more years old, resident in Portugal mainland, using the National Health Service unique identifier User number to link eight electronic health registries. Outcomes included were symptomatic SARS-CoV-2 infections, COVID-19 related hospitalizations or deaths. The exposures of interest were the mRNA vaccines (Cominarty or Spikevax) and the viral vector Vaxzevria vaccine. Complete scheme vaccine effectiveness (VE) was estimated as one minus the confounder adjusted hazard ratio, for each outcome, estimated by time-dependent Cox regression with time dependent vaccine exposure.

**Results:** For the cohort of individuals aged 65-79 years, complete scheme VE against symptomatic infection varied between 43% (Vaxzevria) and 65% (mRNA vaccines). This estimate was slightly lower in the ≥80 year cohort (53% for mRNA vaccines. VE against COVID-19 hospitalization varied between 89% (95%CI: 52-94) for Vaxzevria and 95% (95%CI: 93-97) for mRNA vaccines for the cohort aged 65-79 years and was 76% (95%CI: 67-83) for mRNA vaccines in the ≥80 year cohort. High VE against COVID-19 related deaths were estimated, for both vaccine types, 95% and 81% for the 65-79 years and the ≥80 year cohort, respectively.

We observed a significant waning of VE against symptomatic infection, with VE estimates reaching approximately 34% for both vaccine types and cohorts. Significant waning was observed for the COVID-19 hospitalizations in the ≥80 year cohort (decay from 83% 14-41 days to 63% 124 days after mRNA second dose). No significant waning effect was observed for COVID-19 related deaths in the period of follow-up of either cohorts.

**Conclusions:** In a population with a high risk of SARS-CoV-2 complications, we observed higher overall VE estimates against more severe outcomes for both age cohorts when compared to symptomatic infections. Considering the analysis of VE according to time since complete vaccination, the results showed a waning effect for both age cohorts in symptomatic infection and COVID-19 hospitalization for the 80 and more yo cohort.

## Introduction

Vaccination is an important tool to control COVID-19 pandemic. In Portugal COVID-19 vaccines started to be deployed on December 27, 2020, prioritising individuals with a higher risk of exposure and/or of COVID-19 complications [1]. Since February 3, 2021 a mass vaccination was implemented starting with those aged 80 or more and then by decreasing age groups [1]. The first vaccine to be approved and administered was mRNA Pfizer/BioNTech (Comirnaty) (21/12/2020), followed by Moderna (Spikevax) (06/01/2021), AstraZeneca (Vaxzevria) (29/01/2021) and Janssen vaccine (COVID-19 Vaccine Janssen) (11/03/2021) [2].

The Vaxzevria vaccine was first given to those aged less than 65 years [3], but the recommendation changed in April 2021, to be administrated to those aged more than 60 years old [4]. A single-dose Jansen vaccine was recommended to all males aged 18 and more years old and females aged 50 or more years [5]. In the beginning of October 2021 vaccine coverage in the Portuguese population reached a milestone of 84% (complete scheme) and was estimated to be higher than 95% on those aged 65 and more years [6].

Coinciding with the start of the vaccination campaign, Portugal faced the third and most severe wave of COVID-19. In January 2021, the 14-day incidence rate reached the peak of 1,667 cases per 100,000 inhabitants and the transmissibility (Rt) was over 1 [7]. Given the health care rupture, and to control the epidemic, a lockdown was implemented in January 15^th^, 2021. At this point, the predominant circulating variant was the B.1.7.1 (alpha variant), which was progressively replaced by the B.1.617.2 (delta variant) that became predominant in June 2021. Data on the variants of concern (VOC) surveillance system indicate that, by the end of May 2021 the proportion of circulating variants were 87.7% and 4.8 %, for alpha and delta variants respectively [8]. By the end of June 2021, these figures were 9.8 % and 88.2 %, for alpha and delta variants, respectively [9]. Along with the end of the lockdown and the increasing frequency of the Delta variant, an increasing trend of the epidemic activity was observed in all Portuguese regions, with higher intensity in Algarve and Lisbon and Tagus Valley regions, affecting mainly young adults and adolescents [7]. There were several changes in the national testing strategy along time, but until 13rd of October 2021 the recommendation was still in place to test vaccinated persons in the same circumstances as non-vaccinated (suspected or contact with COVID-19, regular testing in high risk of exposure or high-risk populations, regular testing in specific settings such schools and workplaces).

Post-licencing COVID-19 vaccine studies are needed to measure the protection provided by vaccination in real-world conditions. They add to information provided by clinical trials, as larger and more heterogeneous populations are included. Early evaluations focused mainly on the mRNA Pfizer/BioNTech, and AstraZeneca adenoviral vaccines performance, showing high short-term effectiveness against SARS-CoV-2 infections and severe outcomes in general population and in older adults [10,11]. Vaccine effectiveness (VE) against COVID-19 related hospital admissions and mortality in older adults populations varied between 94-97% for the completed vaccination scheme in December 2020 - April 2021 [10–13]. Following the emergence of the Delta SARS-CoV-2 VOC, studies showed a decrease in VE for both mRNA and AstraZeneca vaccines for symptomatic infections in the general population [14–16], but sustained VE estimates against hospitalizations [14–17] and COVID-19-related mortality [15].

Immunological studies suggest a decrease of IgG and neutralizing antibody titres 3-6 months after complete the two-dose schedule [18,19]. Continued monitoring of vaccine effectiveness over time is important to provide inputs of a potential vaccine waning effect, as has been observed with vaccines against other respiratory pathogens [20,21] and COVID-19 vaccines [16].

In a context of multiple vaccines and variant epidemiological setting, it is important to access the protection conferred by the COVID-19 vaccines in use. Moreover, monitoring over time allows detection of potential decay of vaccine effectiveness and may be important for decision on the vaccination strategy. This study aimed to estimate COVID-19 vaccines effectiveness against symptomatic infections, COVID-19-related hospitalizations and deaths in Portuguese adults aged 65 years old or more in February-September 2021.

## Methods

### Study design

An historical cohort study, based on deterministic data linkage of nationwide electronic health registries [15] was designed to measure VE of COVID-19 vaccines used in Portugal according to time since exposure, against three outcomes: i) symptomatic infection, ii) COVID-19 hospitalizations and iii) COVID-19 related deaths in individuals aged 65 or more years old (yo).

Data extraction and linkage were performed on 16 September 2021 by the Shared Services of the Portuguese Ministry of Health. The National Health Service User (NHSU) unique numeric identifier was used to link all used databases. These included the NHSU, the national vaccination registry, the National Information System for Epidemiologic Surveillance, the National Death Registry, the Primary Information System, the Primary Care Clinical Monitoring System of COVID-19 Patients in Home Isolated, the National Database of Medicine and Treatment Prescriptions and the National Database of Hospital Discharges.

#### Target population and exclusion criteria

The study population comprised Portuguese residents aged 65 or more years old registered in the NHSU database and residing in Portugal mainland, eligible for COVID-19 vaccination during the study period.

Individuals resident in long-term care facilities and other institutions were excluded as timing for vaccination and potential exposure to SARS-CoV-2 were different from community dwelling individuals. Given the use of electronic registries, that does not allow proper identification of institutionalized individuals, the cohort was set when the vaccination coverage in the population considered a priority for vaccination, which also includes the population living in long-term facilities, was higher than 80%.

The study was also restricted to those aged less than 110 years and to those with no prior SARS-CoV-2 infection. For those aged 65 to 79 years old, to ensure that the cohort included individuals with the same likelihood to be vaccinated, only the subset of frequent user of the national health service [22], i.e., individuals who had at least one contact with a primary health care unit in the previous 3 years, were included in the analysis. Finally, for the 80 and more years population, the cohort was also restricted to those who received at least one influenza or pneumococcal vaccine in the last 5 years to increase the likelihood of being a current health care user.

#### Study Period

The observation period was established for each age-group cohort in accordance with the vaccination plan rollout. Namely, from 2nd February 2021 for individuals with 80 or more years and 30th of March 2021 for the 65 to 79 years of age, up to the date of the last observed event in each cohort (date of last observed event in supplementary material).

#### Variables

##### Outcomes

An individual was considered as having had a symptomatic infection if she/he had a laboratory confirmed infection and at least one of the following symptoms: feverishness/ fever, cough, sore throat, dyspnea, anosmia, diarrhea, abdominal pain, fatigue, malaise, myalgia, nausea or vomiting in an interval of <25 to >15 days since laboratory confirmation for SARS-CoV-2 infection notified in the National Information System for Epidemiologic Surveillance or in the Primary Care Clinical Monitoring System of COVID-19 Patients in Home Isolated.

In Portugal, any individual presenting symptoms suspicious of SARS-CoV-2 infection or that was a contact of an infected individual is tested by Nucleic Acid Amplification Tests (TAAN) or Rapid Antigen Test (RAT) if TAAN were not available. Also, all persons in high risk of exposure (eg. Health Care Workers), or in high risk of complications or moving into specific settings (long term care facilities, schools, workplaces with more than 150 professionals) should be regularly tested regardless of their vaccination status until 13 of October 2021. TAAN tests should be used in high risk contexts, and RAT are accepted in other settings [23]. Nevertheless, vaccinated persons might be less prone to accept being regularly tested as their risk perception decreases after vaccination which might reduce the probability to find asymptomatic infected people within the vaccinated population when compared to those not vaccinated.

For COVID-19 hospitalization, we considered all individuals with a date of laboratory confirmation within the study period and hospital discharge with COVID-19 as primary diagnosis (ICD10 code U07.1) [24]. Hospitalizations were retrieved through the national database of hospital discharges that contains all hospitalizations discharge from public hospitals in mainland Portugal. After discharge, all hospitalizations are coded by trained professionals according to the international classification of disease (ICD), 10^th^ version. Timeliness of codification may vary between regions and hospitals, and may take up to 6 months for completion.

A COVID-19 related death was defined as all-cause death with a positive RT-PCR test within the previous 30 days [25].

##### Exposure

Data on vaccine uptake were collected from the national vaccination registry. Complete vaccination was considered 14 or more days after the second dose administration (for vaccines with a two-dose scheme). To evaluate the hypothesis of vaccine effectiveness waning, time after complete vaccination was stratified in 28-day intervals.

##### Confounding factors

Potential confounders included age groups, sex, municipality level European Deprivation Index (EDI) quintile [26], number of chronic diseases, number of laboratory SARS-CoV-2 tests during 2020, previous influenza or pneumococcal vaccines uptake. Demographic data was collected from the NHU database, EDI by municipalities were previously calculated [26], chronic diseases considered were collected from the primary care database (anemia, asthma, cancer, cardiac disease, stroke, dementia, diabetes, hypertension, chronic liver disease, neuromuscular disease, renal disease, rheumatologic disease pulmonary disease, obesity, immunodeficiency, tuberculosis), SARS-CoV-2 tests were extracted from the National Information System for Epidemiologic Surveillance and other vaccines from national vaccination registry.

## Statistical methods

Descriptive statistics were used to characterize study participants. VE was estimated by vaccine type (mRNA or Vaxzevria). Given the different vaccination and observational periods, we estimated vaccine effectiveness (VE) separately for each age-group cohort 65-79 and ≥80 years of age.

Observation time was stratified in 7-day intervals from the start to the ending date of the observational periods, in each cohort. Time dependent person-year exposure experience was calculated for unvaccinated and vaccinated persons according to the number of doses and time of exposure to each dose. Participants who developed one primary outcome or who died during the study period were excluded from the denominator after the event.

For each outcome, incidence rates per 100,000 person years were calculated for unvaccinated exposure and for vaccine exposure period, according to the number of doses and time since each dose. Crude incidence rate ratios among each vaccination group and unvaccinated were calculated with their respective 95% confidence intervals.

VE was computed as one minus the confounder adjusted hazard ratio, for each outcome, estimated by time-dependent Cox regression, adjusted for age group, sex, municipality level EDI quintile, number of chronic conditions, number of SARS-CoV-2 tests in 2020 and uptake of influenza or pneumococcal vaccine in previous 3 years of study period.

The hypothesis of VE waning with time since complete vaccination was evaluated by comparing post-estimation Cox regression hazard ratio of each outcome between the group of individuals with 14 to 41 days of exposure to those with, 124 or more days for ≥80 cohort group,. For 65- 79 due to shorter follow up period VE waning hypothesis was assessed for 98+ days and 70+ days since complete vaccination for symptomatic infection and for both severe outcomes, respectively.. All statistical analysis was performed in R Computing Environment, version 4.0.5.

### Ethical considerations

The study protocol was approved by the Ethical Committee of the Instituto Nacional de Saúde Doutor Ricardo Jorge (INSA) and the data protection officers of INSA, General Directorate for Health, ACSS and SPMS. All data were anonymized prior to analysis.

## Results

### Participants

Database extraction at 16^th^ September 2021 contained 2,117,002 individuals aged 65 to 79 years and 923,450 individuals aged ≥80 years. After excluding, according to previously described criteria, the cohorts were constituted, by 1,414,909 aged 65 to 79 years and 470,023 aged ≥80 years cohort (Flowchart in Supplementary material).

Distribution of vaccines varied by age cohort. Namely, for the 65 to 79 yo cohort, the Pfizer/BioNTech vaccine was administrated to 644,959 individuals (45.6%) and the second most frequent vaccine was from AstraZeneca (499,770 persons, 35.3%). Vaccines from Moderna (115,217, 8.1%) and Jassen (40,484, 2.9%) were less frequently administered in this age cohort. The majority of adults aged 80 and more yo, were vaccinated with the Pfizer/BioNTech vaccine (379,285, 80.7%). The other vaccines were also available for this age group, but were less frequently used (Moderna: 56,037 individuals, 11.9%; AstraZeneca: 8,509 individuals, 1.8%; Jassen: 2,191 individuals, 0.5%) (Flowchart in Supplementary material).

Comparing the vaccinated with the unvaccinated group, we observed some differences in age structure and number of chronic conditions. The unvaccinated were older then vaccinated with AstraZeneca in the 80 and more yo cohort (median of 86 vs 82, respectively). In both cohorts, the proportion of individuals without any chronic condition was higher in the unvaccinated group than in the vaccinated with AstraZeneca or mRNA vaccines (65 to 79 yo cohort: 48.9% vs 22.9% or 29.3% or 22.7%; 80 and more yo cohort: 37.2% vs 11.2% or 14.3% or 10.2%) (Table S1 and Table S2, supplementary material).

During the study period, in the 65 to 79 years cohort, a total of 4,282 symptomatic infections, 338 hospitalizations and 172 deaths were observed (Table 2), compared to a total of 3,078 symptomatic infections, 854 hospitalizations and 764 COVID-19 related deaths observed in the 80 and more yo cohort(Table 3).

## Main results

### Vaccine effectiveness against symptomatic infection

For mRNA VE against symptomatic infection varied between 53% (95%CI: 45 to 60) in the 80 and more yo and 65% (95%CI: 62 to 68) in the 65-79 yo cohort. Considering the AstraZeneca vaccine, VE estimate was 43% (95%CI: 37 to 49) in the 65-79 yo cohort

Analysis of vaccine effectiveness according to time since complete vaccination showed a reduction for mRNA vaccines. The reduction varied according to the age cohort in analysis, potentially related to the different time of observation for each cohort. Decline was observed in the 80 and more yo cohort, (hazard ratio of symptomatic infection of 2.33), with VE estimates decreasing from 72% (14 to 41 days after complete vaccination) to 34% (124 to 203 days after complete vaccination). For the cohort of individuals aged between 65 to 79 years, the VE estimates declined from 79% at 14 to 41 days after the second dose, to 39% at 98 and more days since second dose, corresponding to a hazard ratio of 2.9 (2.4 to 3.6) (Table 1)

**Table 1:** Symptomatic SARS-CoV-2 infection, COVID-19 related hospitalizations and deaths, incidence, hazard ratios and vaccine effectiveness by mRNA (Cominarty or Spikevax) and Vaxzevria vaccination status for individuals aged 65-79 years, Portugal, March–September 2021 (n = 1,414,909)

| Vaccine Status | Person<br>years | events<br>(n) | Rate per<br>10,000 py | Confounder-adjusted HR 95% CI | VE 95% CI |
| --- | --- | --- | --- | --- | --- |
| **Symptomatic SARS-CoV-2 infection** |  |  |  |  |  |
| mRNA vaccine (Cominarty or Spikevax) - Total | 361,642 | 3,185 |  |  |  |
| Unvaccinated | 156,266 | 1,457 | 93.2 |  |  |
| **Complete vaccination** | **205,376** | **1,728** | **84.1** | **0.35 (0.32 to 0.38)** | **65 (62 to 68)** |
| **14-41 days** | **57,355** | **188** | **32.8** | **0.21 (0.17 to 0.24)** | **79 (76 to 83)** |
| 42-69 days | 56,258 | 521 | 92.6 | 0.32 (0.29 to 0.36) | 68 (64 to 71) |
| 70-97 days | 53,145 | 596 | 112.1 | 0.41 (0.37 to 0.47) | 59 (53 to 63) |
| **≥98 days (max=148)** | **38,618** | **423** | **109.5** | **0.61 (0.52 to 0.71)** | **39 (29 to 48)** |
| *Waining effect* |  |  |  | 2.9 (2.4 to 3.6) |  |
| Vaxzevria – Total | 233,648 | 2,554 |  |  |  |
| Unvaccinated | 156,266 | 1,457 | 93.2 |  |  |
| **Complete vaccination** | **77,386** | **1,097** | **141.8** | **0.57 (0.51 to 0.63)** | **43 (37 to 49)** |
| **14-41 days** | **37,135** | **546** | **147.0** | **0.52 (0.46 to 0.58)** | **48 (42 to 54)** |
| 42-69 days | 33,830 | 495 | 146.3 | 0.67 (0.58 to 0.77) | 33 (23 to 42) |
| **≥70 days (max=142)** | **6,417** | **56** | **87.3** | **0.66 (0.48 to 0.90)** | **34 (10 to 52)** |
| *Waining effect* |  |  |  | 1.27 (0.92 to 1.76) |  |
| **COVID-19 related hospitalizations** |  |  |  |  |  |
| mRNA vaccine (Cominarty or Spikevax) – Total | 362,147 | 298 |  |  |  |
| Unvaccinated | 156,564 | 266 | 17.0 |  |  |
| **Complete vaccination** | **205,583** | **32** | **1.6** | **0.05 (0.03 to 0.07)** | **95 (93 to 97)** |
| **14-41 days** | **57,366** | **10** | **1.7** | **0.05 (0.03 to 0.90)** | **95 (90 to 97)** |
| 42-69 days | 56,293 | 9 | 1.6 | 0.03 (0.02 to 0.06) | 97 (94 to 98) |
| **≥70 days (max=148)** | **91,924** | **13** | **1.4** | **0.07 (0.04 to 0.14)** | **93 (86 to 96)** |
| *Waining effect* |  |  |  | 1.35 (0.57 to 3.22) |  |
| Vaxzevria - Total | 234,086 | 286 |  |  |  |
| Unvaccinated | 156,564 | 266 | 17.0 |  |  |
| **Complete vaccination** | **77,522** | **20** | **2.6** | **0.11 (0.06 to 0.48)** | **89 (52 to 94)** |
| **COVID-19 related deaths** |  |  |  |  |  |
| mRNA vaccine (Cominarty or Spikevax) - Total | 362,210 | 159 |  |  |  |
| Unvaccinated | 156,623 | 129 | 8.2 |  |  |
| **Complete vaccination** | **205,587** | **30** | **1.5** | **0.05 (0.03 to 0.08)** | **95 (92 to 97)** |
| **14-41 days** | **57,366** | **5** | **0.9** | **0.05 (0.02 to 0.12)** | **95 (88 to 98)** |
| 42-69 days | 56,294 | 7 | 1.2 | 0.03 (0.02 to 0.08) | 97 (92 to 98) |
| **≥70 days (max=148)** | **91,927** | **18** | **2.0** | **0.07 (0.04 to 0.13)** | **93 (87 to 96)** |
| *Waining effect* |  |  |  | 1.34 (0.48 to 3.75) |  |
| Vaxzevria - Total |  |  |  |  |  |
| Unvaccinated | 156,623 | 129 | 8.2 |  |  |
| **Complete vaccination** | **77,527** | **13** | **1.7** | **0.05 (0.03 to 0.10)** | **95 (90 to 97)** |

**SARS-CoV-2 symptomatic case:** individual with a SARS-CoV-2 laboratory confirmation during study period and at least one symptom (feverishness/ fever, cough, sore throat, dyspnea, anosmia, diarrhea, abdominal pain, fatigue, malaise, myalgia, nausea or vomiting) declared from in the period -15 to 25 days of laboratory confirmation; Complete vaccination: 2 doses ≥ 14 days; Rate: per 1,000 person-years; Confounder-adjusted HR: confounder-adjusted hazard ratio obtained by time-dependent Cox regression with vaccine exposure as time-dependent, adjusted for age group, sex, health region, municipality level European Deprivation quintiles, number of chronic diseases, number of SARS-CoV-2 tests performed in 2021, influenza or pneumococcal vaccine uptake in the past 3 years and time (7-day periods); VE was calculated by $(1 - HR) \times 100$ .

**Table 2:**
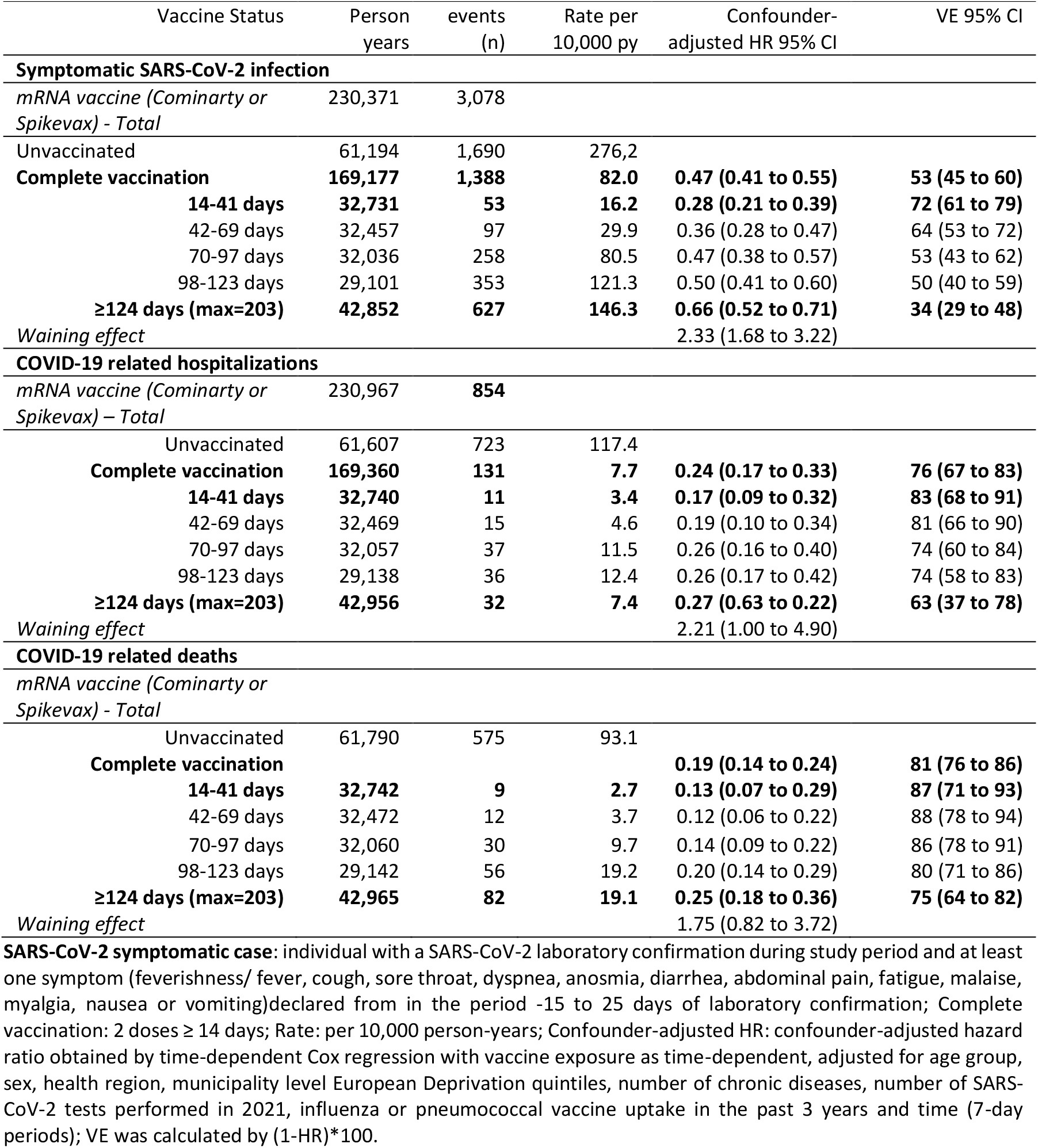
Symptomatic SARS-CoV-2 infection, COVID-19 related hospitalizations and deaths, incidence, hazard ratios and vaccine effectiveness by mRNA (Cominarty or Spikevax) vaccination status for individuals aged ≥80 years, Portugal, February–September 2021 (n = 470,023)

Although not significant, we also observed reduction of VE in the AstraZeneca vaccines for the 65-79 yo cohort, for whom this vaccine was more frequently used and with sufficient time of observation. VE reduced from 48% (14 to 41 days from complete vaccination) to 34% (70 or more days from complete vaccination). Hazard ratio between the two periods was estimated in 1.27 (95%CI: 0.92 to 1.76).

### Vaccine effectiveness against hospitalizations

Vaccine effectiveness against more severe outcomes were higher when compared to symptomatic infection. For the 65-79 yo cohort VE varied between 89% (AstraZeneca vaccines) and 95% (mRNA vaccines). For the 80 and more yo cohort, mRNA VE against hospitalizations was estimated at 76%. Due to small size of the group exposed among 80 or more years old population VE against hospitalizations for AstraZeneca vaccine was not estimated.

Considering time since vaccination, for the 80 years of age or more cohort, mRNA VE against COVID-19 hospitalization was 83% (14 to 41 days after complete vaccination) and 63% (124 or more days from complete vaccination). These estimates translated into a decrease of protection between the two periods, with a 2.21 (95% CI 1.0 to 4.9) hazard rate ratio of COVID-19-related hospitalization.

### Vaccine effectiveness against COVID-19 related deaths

High vaccine effectiveness were estimated for both age cohorts and vaccines. For the mRNA vaccines, estimates varied between 81% (80 and more yo cohort) and 95% (65- to 79 yo cohort). For the AstraZeneca vaccine, VE against COVID-19 related deaths was estimated at 95% in the 65-79 years old group.

Although non-significant, the results showed a reduction in mRNA VE estimates with time in the older cohort. In the 80 and more yo cohort, VE varied from 87% to 75%, with a hazard ratio of 1.75 (95 % CI: (0.82 to 3.72) between 124 or more days versus 14 to 41 days after the second dose.

## Discussion

### Key results

In this study we present VE estimates against SARS-CoV-2 symptomatic infection, COVID-19 hospitalization and COVID-19 related death for the mainland Portuguese population aged 65 and more years. Main results indicate that VE varied according to the outcome and vaccine type. VE estimates against symptomatic infections (vaccine type specific VE range from 43% to 65% in the 65-79 yo cohort and 53% in the 80 and more yo cohort). These estimates were lower than VE estimated against severe COVID-19 outcomes – hospitalization and death (higher than 89% in the 65-79 yo cohort and higher than 76% in the 80 and more yo cohort). These results are indicative of a high level of vaccine protection against severe COVID-19 outcomes in the 65 and more yo population that is more prone to COVID-19 related complications.

Compared to other studies we found some differences. A living systematic review that included studies published (in preprint) up to 14 May 2021, indicated that VE against all infections (symptomatic and asymptomatic) ranged between 61.7% e 98.6%, depending on the age group and vaccine. An update of this living review, that included studies published up to 25 August 2021, allowed a meta-analysis and indicated that VE against symptomatic infection was 75.7% (95%CI: 69.3 to 80.8) [27]. The differences between our overall VE estimates and other studies might be explained by different study periods and consequently different epidemiological and virological contexts.

Our study comprise a period of high incidence observed at the beginning of the vaccination campaign, corresponding to the third COVID-19 peak in January-February 2021 (for the 80+ cohort), but also the fourth wave associated with the circulation of SARS-CoV-2 Delta variant during May-September 2021. As described in the living review, when estimating VE comparing Delta vs Alpha, estimates tended to be 10 to 20% lower for less severe COVID-19 infection but not different for severe outcomes [27].

Considering VE against COVID-19 related death, we could find less studies that focused on this severe outcome, as the majority reported VE against severe outcomes which included hospitalizations or death and thus are not comparable with our results [14,25]. Results from one study conducted in the population aged 60 or more years in Scotland reported high VE against death (defined as death with Covid-19 recorded in the death certificate or that occurred within 28 days after a positive RT-PCR test) of 90% (95% CI: 84 to 94) for AstraZeneca vaccine and 87% (95% CI: 77 to 93) for Pfizer vaccine [28]. Our VE estimates against death are in line with these estimates and also support a high protection conferred by both vaccines when considering this fatal outcome.

For the 60-79 yo cohort, we were able to estimate VE for the different outcomes and for two types of vaccines: mRNA (Cominarty and Spikevax) and adenovirus based vaccine (Vaxzevria). Similar analysis was not possible for the 80 and more cohort or for the Jassen vaccine, as the proportion of vaccinated individuals was too low. For the 65-79 yo cohort and considering symptomatic infection, VE estimates indicate a lower protection conferred by the adenovirus vaccine [Vaxzveria VE of 43%; 95%CI 37 to 49 vs mRNA (Cominarty or Spikevax) VE of 65% with 95%CI of 62 to 68]. Other brand or type specific vaccine studies have also reported similar lower VE against symptomatic infection of AstraZeneca when compared to Pfizer vaccine [29,30].

After completed vaccination schemes, we observed a decay of the VE estimates with time, particularly against symptomatic infection and in the 80 and more yo cohort, where VE decay was observed for both symptomatic infection and hospitalizations. The drop over time of VE against symptomatic infection has also been reported by several authors [14–16] but the decay of VE against hospitalization had not yet observed as far as we know [14–17]. In part, this result may be explained by an insufficient time of follow up. Our results indicate a significant decay of VE against hospitalizations in the 80 and more yo cohort, from 83% (the peak achieved 14-41 days after complete vaccination) to 63% (after 124 days of complete vaccination). One valid argument, is that this decay may be attributable to the increased circulation of the Delta variant which could have increased the effect on the older aged cohort. However waning of vaccine effect with time cannot be ruled out. As observed by Tartof et al. [16], the decay on VE against symptomatic infection was observed on all population (even in younger aged cohorts with less than <45 yo) and this pattern was observed even before the circulation of Delta variant.

The present study has some limitations, namely related to the datasets used and their quality. For instance, the main dataset used to link data was the NHSU, which contains the unique health number attributed to each individual residing in Portugal. However, as referred, the NHSU database could have update issues, and occasional and temporary registrations of NHS users, such as immigrants and asylum seekers, that could artificially increase the number of individuals in a given cohort. To overcome this limitation, several restriction criteria were included, and from the initial extraction a total of 1,178,975 registries were excluded from the analysis. Final cohort age and sex distribution were comparable to National Statistics estimates for individuals aged 65 and more years [31]. In terms of prevalence of chronic conditions, final cohorts were also comparable to National estimates (INS 2019).

Another potential limitation is related to information bias. In Portugal, during the study period, testing guidelines were not different for vaccinated and non-vaccinated individuals which may have differentially limited the opportunity of infection ascertainment in vaccinated and unvaccinated. Nevertheless, we cannot rule out the hypothesis that vaccinated individuals could have had a differential (due to acceptance) testing performance if asymptomatic or if they had any contact with COVID-19 case. Under this hypothesis there is a potential bias due to different infection ascertainment in vaccinated versus non vaccinated cohorts, which might have contributed to overestimating VE against infection. However, this potential bias is less probable for severe outcomes such as hospitalization. Nevertheless, when we compared the frequency of testing during 2021 between the groups compared in this study we did not observe relevant differences.

In what concerns the COVID-19 hospitalizations outcome, the delay in the update of information in the registries regarding hospital discharge diagnosis might contribute to underrepresent this specific outcome. Although, we didn’t find any reason for having a different delay in the registry of the discharge information between vaccinated and non-vaccinated, there is a possibility of a differential bias if vaccinated people had a lower hospitalization stay when compared to non-vaccinated. Finally, there is potential for residual confounding bias. Adjustment for confounding was made for several variables, most of them very relevant from a clinical and epidemiological point of view. Nevertheless, residual confounding bias could remain mainly due to the absence of information on non-pharmaceutical protective measures such as use of facemask, hand washing and social distancing, all of which could be associated with both vaccine uptake and risk of infection.

Besides the impossibility of excluding the presence of selection or information bias driven by the health seeking behavior, our results are generally in line with results reported in studies performed elsewhere. Finally, as our cohort comprise all the Portuguese NHS users older adults population, we consider our results to represent a description of the direct protection conferred by the COVID-19 vaccines used within the target population, and a good approach to monitor VE along time and waning of the vaccine protection.

## Data Availability

All data produced in the present study are available upon reasonable request to the authors

## Notes

### Competing Interest Statement

The authors have declared no competing interest.

### Funding Statement

This study did not receive any funding

### Author Declarations

The study protocol was approved by the Ethical Committee of the Instituto Nacional de Saude Doutor Ricardo Jorge (INSA) and the data protection officers of INSA, General Directorate for Health, ACSS and SPMS.

