## Supplementary material for "COVID-19 vaccine effectiveness against laboratory confirmed symptomatic SARS-CoV-2 infection, COVID-19 related hospitalizations and deaths, among individuals aged 65 years or more in Portugal: a cohort study based on data-linkage of national registries February-September 2021": Suplemmental tables

Table S1: Demographic characteristics and vaccine status of cohort individuals aged 65 to 79 years and older, Portugal, March–August 2021 (n = 1,414,909).

| | Vaxzevria 499,770 | Janssen 40,484 | mRNA 760,176 | Unvaccinated 114,479 |
|---|---|---|---|---|
| Age (years), median (IQR) | 71.0 (68.0 - 75.0) | 69.0 (67.0 - 72.0) | 71.0 (68.0 - 75.0) | 71.0 (68.0 - 75.0) |
| Age group, (%) | | | | |
| 65-69 | 178,573 (35.7%) | 21,890 (54.1%) | 297,449 (39.1%) | 43,262 (37.8%) |
| 70-74 | 194,191 (38.9%) | 12,776 (31.6%) | 257,681 (33.9%) | 39,124 (34.2%) |
| 75-79 | 127,006 (25.4%) | 5,818 (14.4%) | 205,046 (27.0%) | 32,093 (28.0%) |
| Sex Male, n (%) | 221,369 (44.3%) | 18,407 (45.5%) | 332,106 (43.7%) | 51,302 (44.8%) |
| Region, n (%) | | | | |
| Alentejo | 27,817 (5.6%) | 2,592 (6.4%) | 35,331 (4.6%) | 4,538 (4.0%) |
| Algarve | 27,567 (5.5%) | 6,318 (15.6%) | 29,480 (3.9%) | 12,502 (10.9%) |
| Centro | 97,820 (19.6%) | 10,582 (26.1%) | 133,943 (17.6%) | 19,034 (16.6%) |
| LVT | 212,153 (42.5%) | 11,646 (28.8%) | 234,669 (30.9%) | 41,354 (36.1%) |
| Norte | 132,245 (26.5%) | 8,864 (21.9%) | 322,412 (42.4%) | 25,612 (22.4%) |
| Missing | 2,168 (0.4%) | 482 (1.2%) | 4,341 (0.6%) | 11,439 (10.0%) |
| EDI Quintile, n (%) | | | | |
| Q1 (least deprived) | 66,389 (13.3%) | 10,092 (24.9%) | 118,691 (15.6%) | 13,898 (12.1%) |
| Q2 | 78,578 (15.7%) | 3,584 (8.9%) | 112,593 (14.8%) | 13,854 (12.1%) |
| Q3 | 74,384 (14.9%) | 3,438 (8.5%) | 111,310 (14.6%) | 13,251 (11.6%) |
| Q4 | 137,234 (27.5%) | 10,097 (24.9%) | 221,903 (29.2%) | 27,751 (24.2%) |
| Q5 (most deprived) | 141,017 (28.2%) | 12,791 (31.6%) | 191,338 (25.2%) | 34,286 (29.9%) |
| Missing | 2,168 (0.4%) | 482 (1.2%) | 4,341 (0.6%) | 11,439 (10.0%) |
| Number of chronic diseases, n (%) | | | | |
| 0 | 114,499 (22.9%) | 11,877 (29.3%) | 172,206 (22.7%) | 56,005 (48.9%) |
| 1 | 130,811 (26.2%) | 10,636 (26.3%) | 199,291 (26.2%) | 24,891 (21.7%) |
| 2 | 123,914 (24.8%) | 9,162 (22.6%) | 187,622 (24.7%) | 17,238 (15.1%) |
| 3 | 79,620 (15.9%) | 5,507 (13.6%) | 121,101 (15.9%) | 9,755 (8.5%) |
| 4 | 34,814 (7.0%) | 2,274 (5.6%) | 54,174 (7.1%) | 4,406 (3.8%) |
| 5+ | 16,112 (3.2%) | 1,028 (2.5%) | 25,782 (3.4%) | 2,184 (1.9%) |
| Number of SARS-CoV-2 tests in 2021, n (%) | | | | |
| 0 | 390,166 (78.1%) | 30,999 (76.6%) | 597,080 (78.5%) | 88,703 (77.5%) |
| 1 | 66,129 (13.2%) | 5,226 (12.9%) | 95,874 (12.6%) | 14,323 (12.5%) |
| 2 | 23,741 (4.8%) | 1,865 (4.6%) | 34,980 (4.6%) | 4,943 (4.3%) |
| 3 | 8,557 (1.7%) | 798 (2.0%) | 12,593 (1.7%) | 2,021 (1.8%) |
| 4-9 | 9,835 (2.0%) | 1,277 (3.2%) | 16,586 (2.2%) | 3,514 (3.1%) |
| 10+ | 1,342 (0.3%) | 319 (0.8%) | 3,063 (0.4%) | 975 (0.9%) |
| Any other vaccine, n (%)† | 325,333 (65.1%) | 18,503 (45.7%) | 497,594 (65.5%) | 21,684 (18.9%) |

† received at least one of the following vaccines since 2016,influenza, pn23, pcv 7, 10 or 13

Table S2: Demographic characteristics and vaccine status of cohort individuals aged ≥80 years and older, Portugal, February–August 2021 (n = 470,025).

| | Vaxzevria<br>8,509 | Janssen<br>2,191 | mRNA<br>435,322 | Unvaccinated<br>24,003 |
|---|---|---|---|---|
| Age (years), median (IQR) | 82.0 (80.0 - 86.0) | 85.0 (82.0 - 88.0) | 84.0 (82.0 - 88.0) | 86.0 (83.0 - 90.0) |
| Age group, (%) | | | | |
| 80-84 | 5,681 (66.8%) | 1,078 (49.2%) | 222,717 (51.2%) | 8,997 (37.5%) |
| 85-89 | 2,022 (23.8%) | 732 (33.4%) | 145,526 (33.4%) | 8,137 (33.9%) |
| 90-94 | 660 (7.8%) | 304 (13.9%) | 54,263 (12.5%) | 4,879 (20.3%) |
| 95+ | 146 (1.7%) | 77 (3.5%) | 12,816 (2.9%) | 1,990 (8.3%) |
| Sex Male, n (%) | 3,517 (41.3%) | 769 (35.1%) | 176,846 (40.6%) | 8,645 (36.0%) |
| Region, n (%) | | | | |
| Alentejo | 1,308 (15.4%) | 186 (8.5%) | 24,081 (5.5%) | 1,106 (4.6%) |
| Algarve | 884 (10.4%) | 313 (14.3%) | 15,897 (3.7%) | 1,412 (5.9%) |
| Centro | 1,547 (18.2%) | 519 (23.7%) | 91,855 (21.1%) | 4,627 (19.3%) |
| LVT | 2,710 (31.8%) | 699 (31.9%) | 142,581 (32.8%) | 7,969 (33.2%) |
| Norte | 2,012 (23.6%) | 460 (21.0%) | 159,442 (36.6%) | 8,095 (33.7%) |
| Missing | 48 (0.6%) | 14 (0.6%) | 1,466 (0.3%) | 794 (3.3%) |
| EDI Quintile, n (%) | | | | |
| Q1 (least deprived) | 1,078 (12.7%) | 366 (16.7%) | 76,030 (17.5%) | 3,836 (16.0%) |
| Q2 | 1,032 (12.1%) | 285 (13.0%) | 68,094 (15.6%) | 3,403 (14.2%) |
| Q3 | 1,112 (13.1%) | 268 (12.2%) | 66,021 (15.2%) | 3,582 (14.9%) |
| Q4 | 2,294 (27.0%) | 519 (23.7%) | 120,749 (27.7%) | 6,312 (26.3%) |
| Q5 (most deprived) | 2,945 (34.6%) | 739 (33.7%) | 102,962 (23.7%) | 6,076 (25.3%) |
| Missing | 48 (0.6%) | 14 (0.6%) | 1,466 (0.3%) | 794 (3.3%) |
| Number of chronic diseases, n (%) | | | | |
| 0 | 956 (11.2%) | 313 (14.3%) | 44,531 (10.2%) | 8,927 (37.2%) |
| 1 | 1,529 (18.0%) | 382 (17.4%) | 82,962 (19.1%) | 3,687 (15.4%) |
| 2 | 2,024 (23.8%) | 523 (23.9%) | 112,314 (25.8%) | 4,258 (17.7%) |
| 3 | 1,885 (22.2%) | 471 (21.5%) | 96,833 (22.2%) | 3,620 (15.1%) |
| 4 | 1,207 (14.2%) | 288 (13.1%) | 58,377 (13.4%) | 2,002 (8.3%) |
| 5+ | 908 (10.7%) | 214 (9.8%) | 40,305 (9.3%) | 1,509 (6.3%) |
| Number of SARS-CoV-2 tests in 2021, n (%) | | | | |
| 0 | 5,651 (66.4%) | 1,330 (60.7%) | 327,670 (75.3%) | 15,614 (65.1%) |
| 1 | 1,304 (15.3%) | 329 (15.0%) | 52,314 (12.0%) | 3,110 (13.0%) |
| 2 | 624 (7.3%) | 165 (7.5%) | 22,336 (5.1%) | 1,719 (7.2%) |
| 3 | 330 (3.9%) | 86 (3.9%) | 11,110 (2.6%) | 1,033 (4.3%) |
| 4-9 | 524 (6.2%) | 230 (10.5%) | 19,170 (4.4%) | 2,218 (9.2%) |
| 10+ | 76 (0.9%) | 51 (2.3%) | 2,722 (0.6%) | 309 (1.3%) |
| Any other vaccine, n (%)† | 8,047 (94.6%) | 1,984 (90.6%) | 420,208 (96.5%) | 19,865 (82.8%) |

† received at least one of the following vaccines since 2016,influenza, pn23, pcv 7, 10 or 13

Graphs

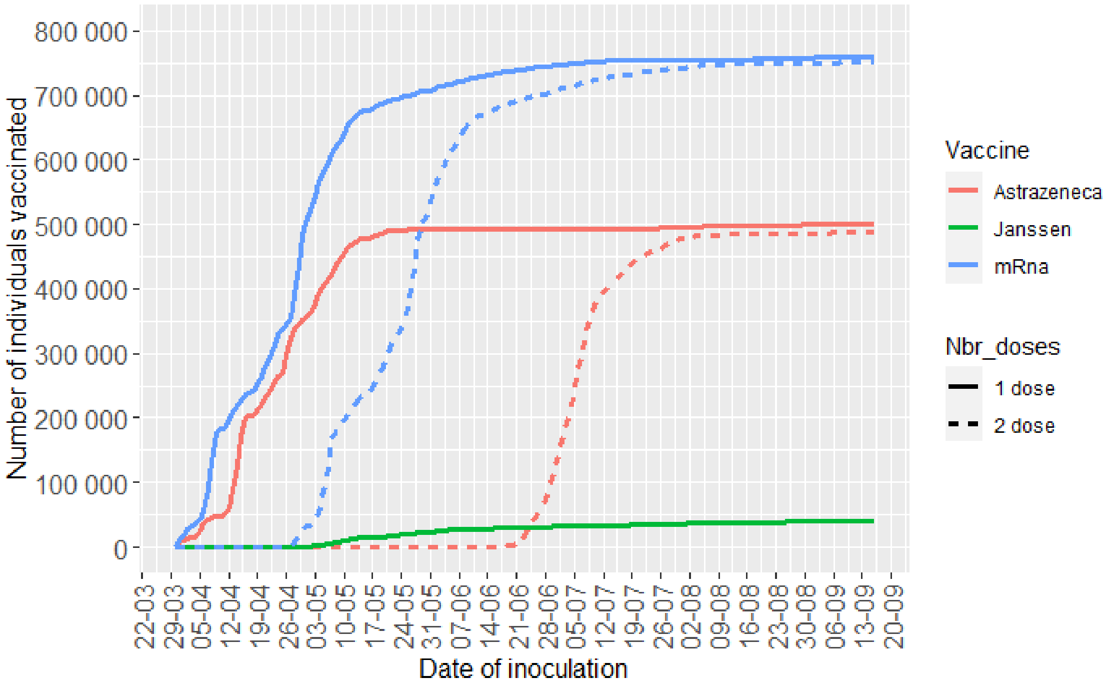


Figure S1: 65-79

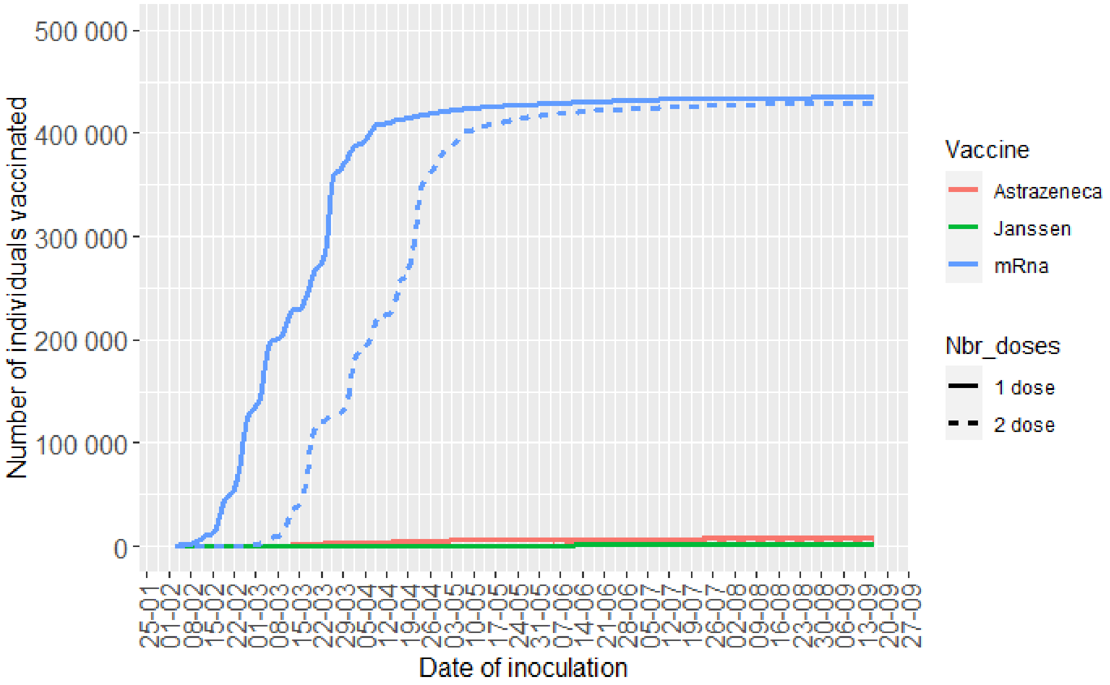


Figure S2 ≥80

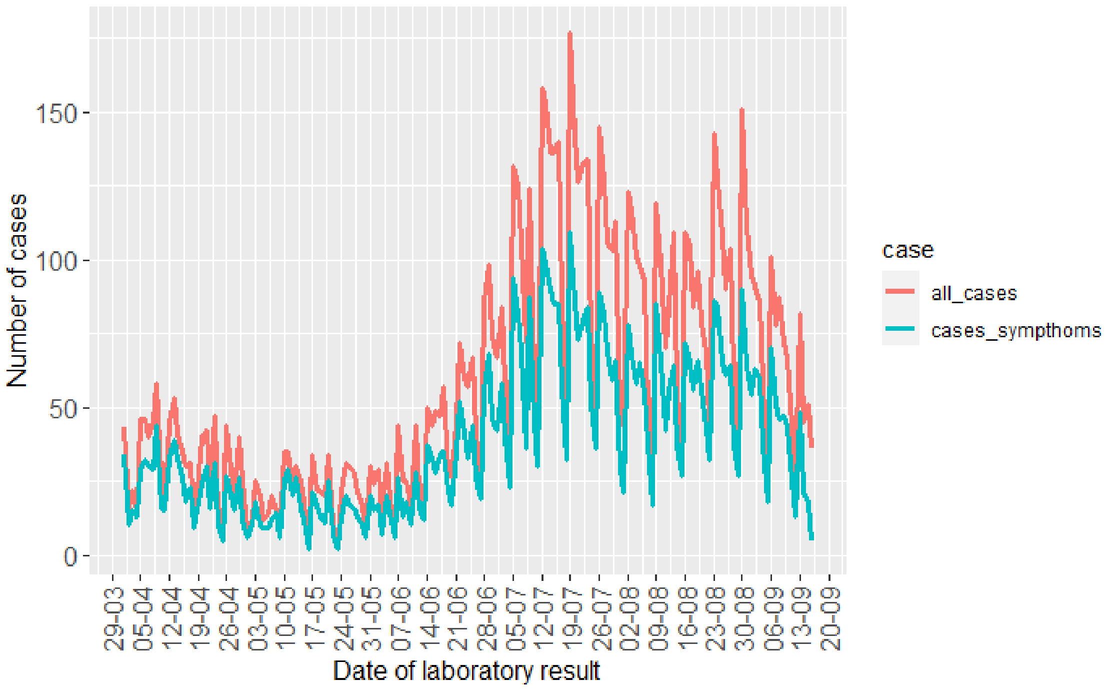


Figure S4 SARS-CoV-2 infection cases in the period of study cohort 65-79 years of age

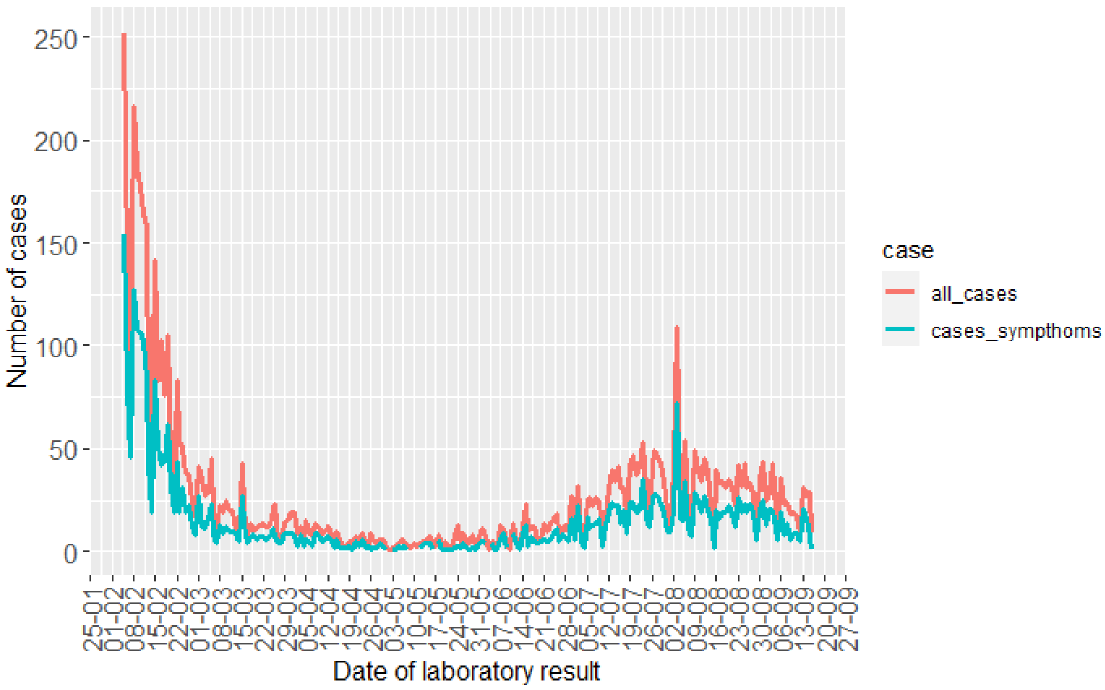


Figure S5 SARS-CoV-2 infection cases in the period of study cohort ≥80 years of age

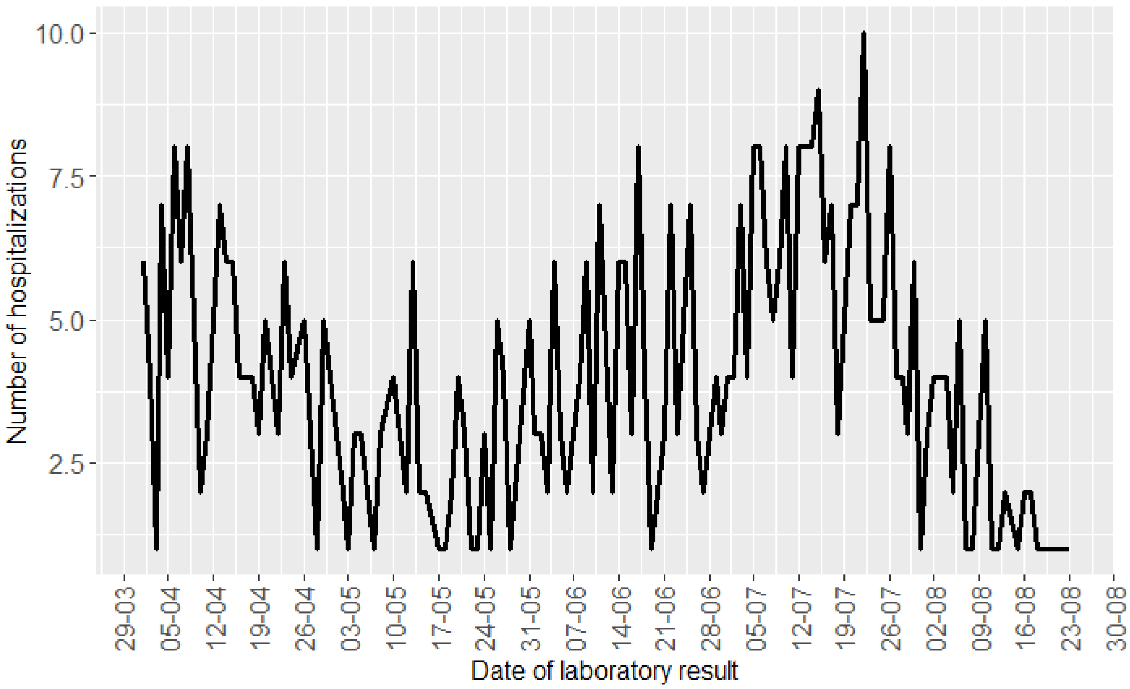


Figure S7 COVID-19 related hospitalizations in the period of study cohort 65-79 years of age

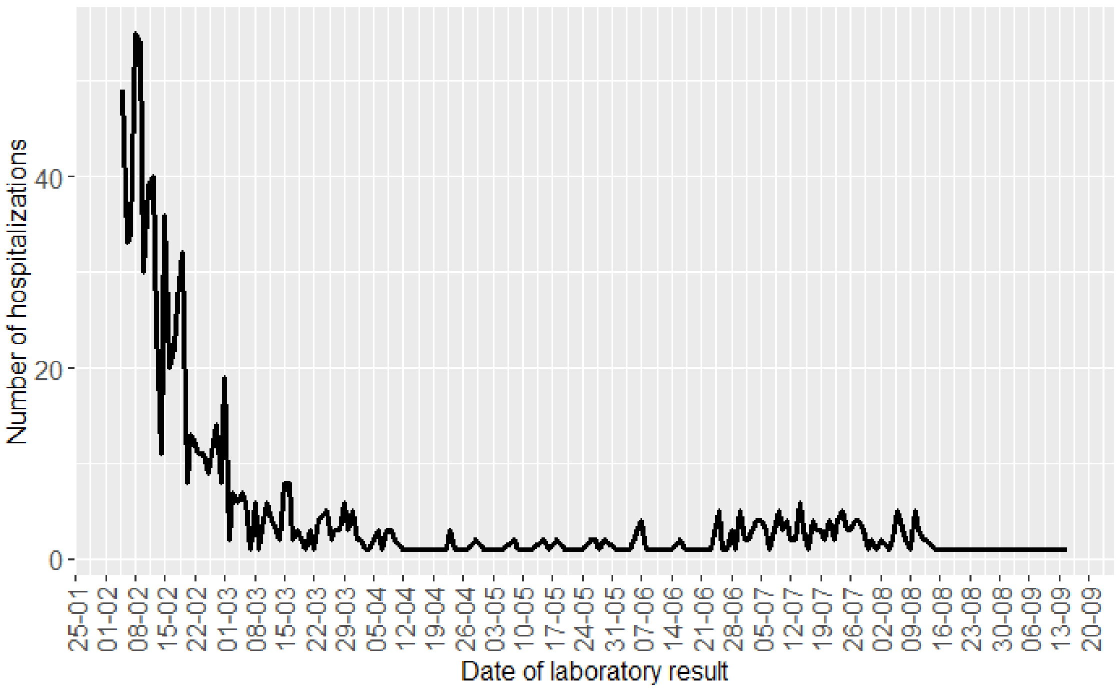


Figure S8 COVID-19 related hospitalizations in the period of study cohort ≥80 years of age

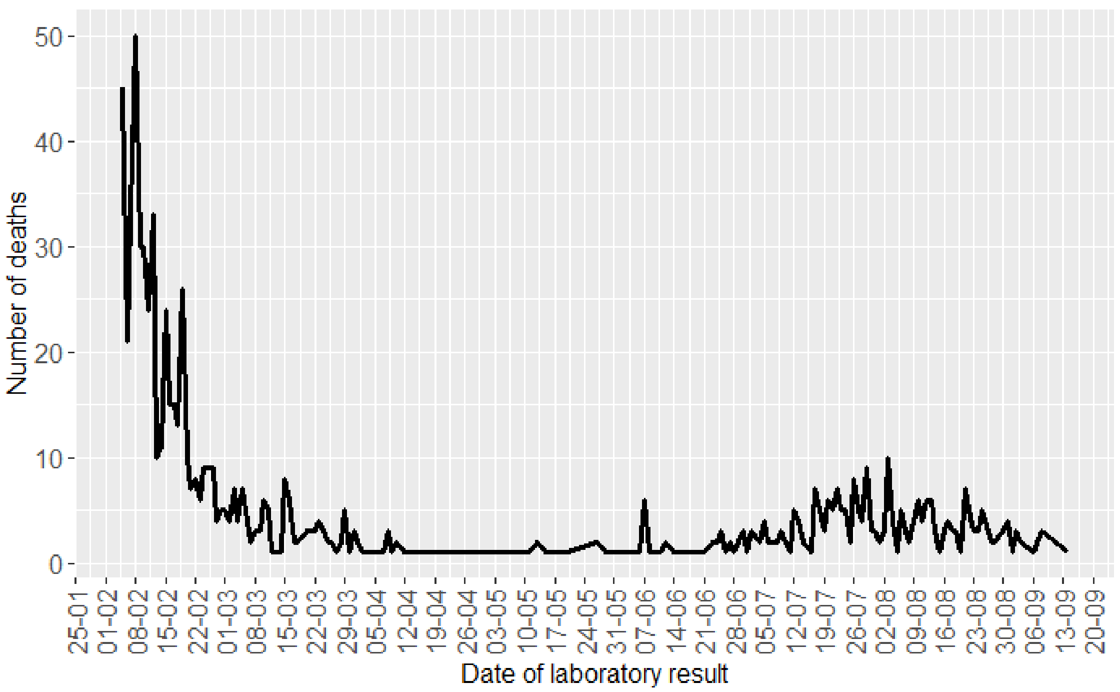


Figure S9 COVID-19 related deaths in the period of study cohort ≥80 years of age

Flowchart

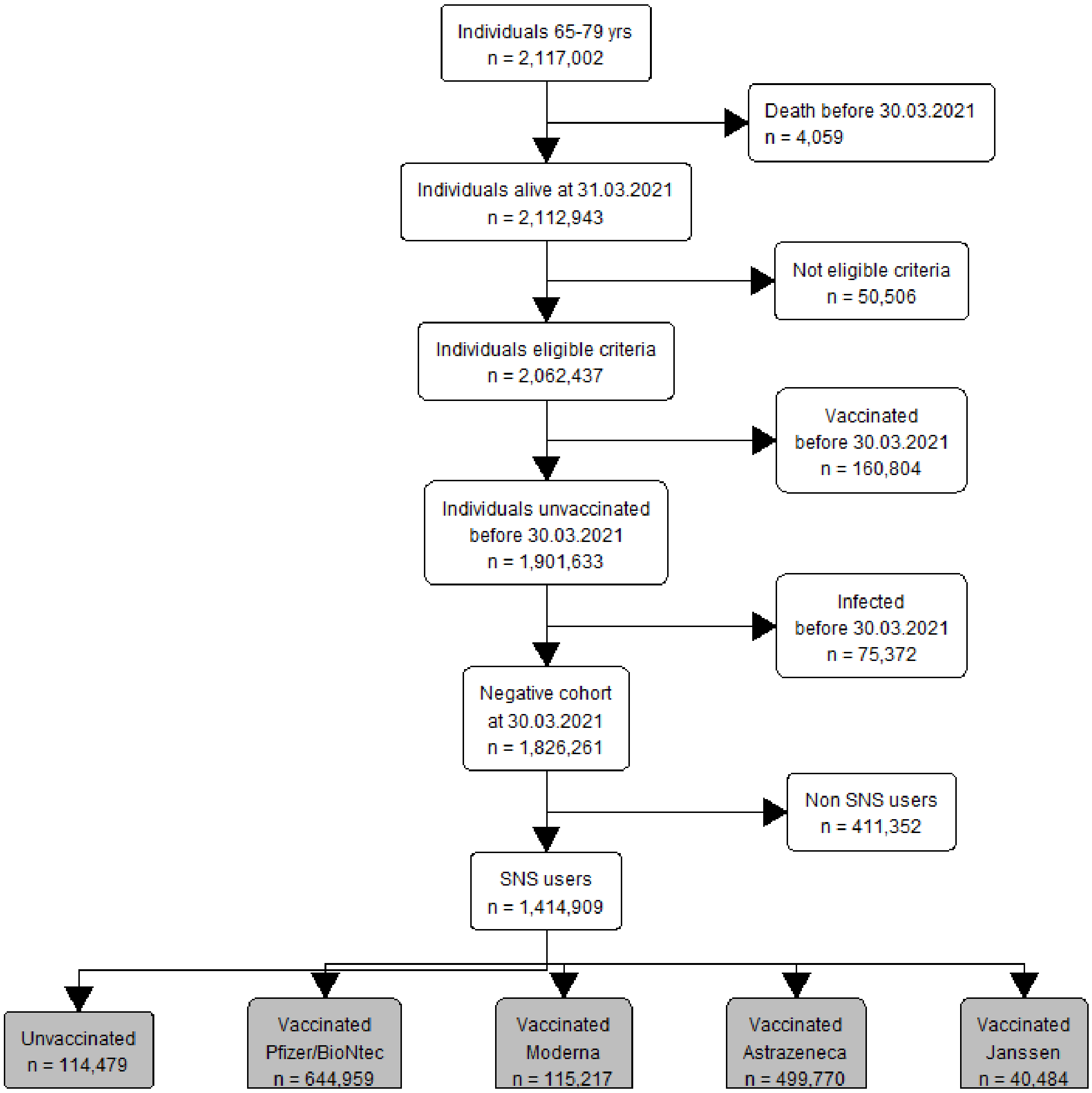
Individuals 65-79 yrs
n = 2,117,002
Death before 30.03.2021
n = 4,059
Individuals alive at 31.03.2021
n = 2,112,943
Not eligible criteria
n = 50,506
Individuals eligible criteria
n = 2,062,437
Vaccinated
before 30.03.2021
n = 160,804
Individuals unvaccinated
before 30.03.2021
n = 1,901,633
Infected
before 30.03.2021
n = 75,372
Negative cohort
at 30.03.2021
n = 1,826,261
Non SNS users
n = 411,352
SNS users
n = 1,414,909
Unvaccinated
n = 114,479
Vaccinated
Pfizer/BioNtec
n = 644,959
Vaccinated
Moderna
n = 115,217
Vaccinated
Astrazeneca
n = 499,770
Vaccinated
Janssen
n = 40,484

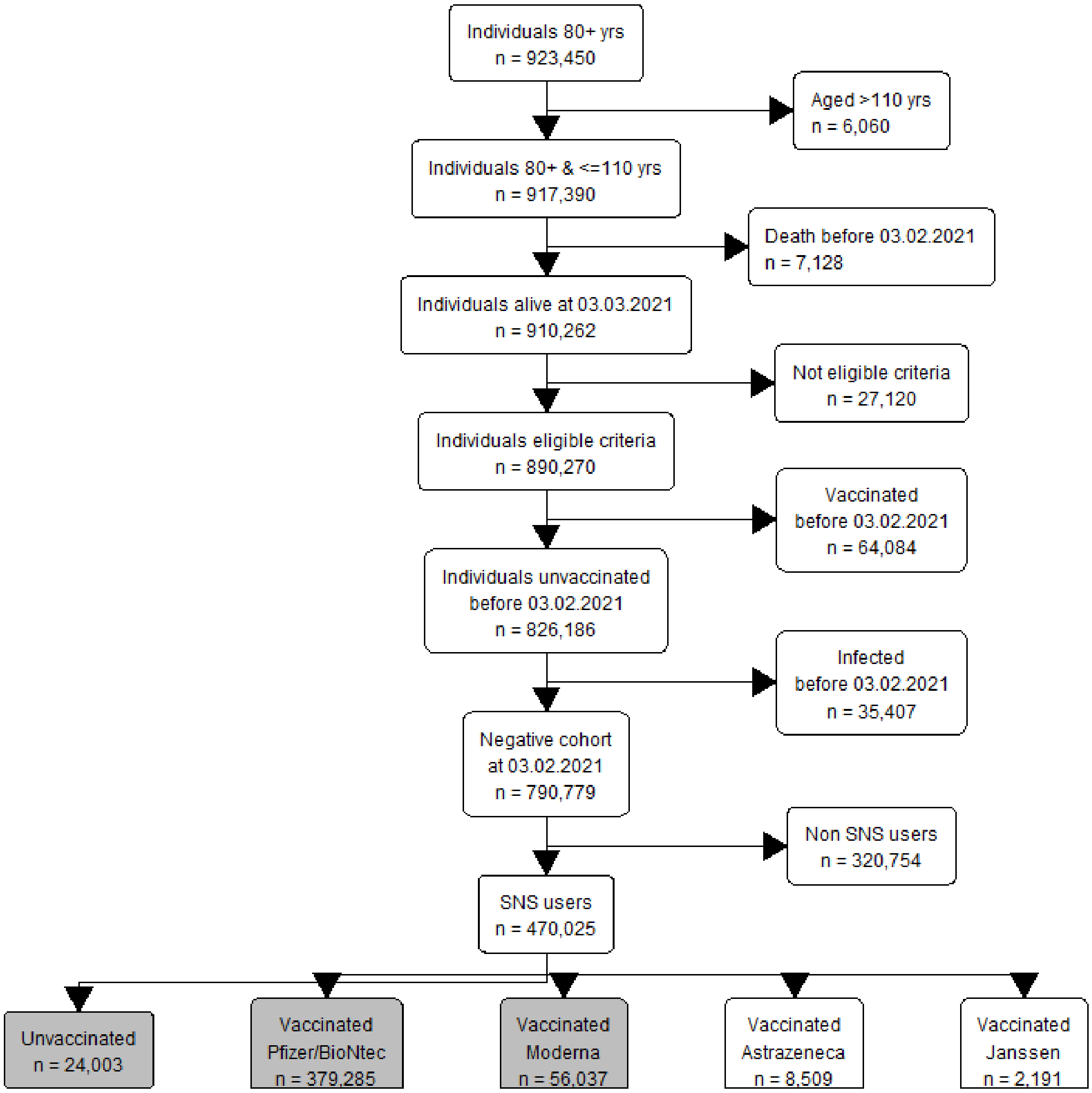
Individuals 80+ yrs
n = 923,450
Aged >110 yrs
n = 6,060
Individuals 80+ & <=110 yrs
n = 917,390
Death before 03.02.2021
n = 7,128
Individuals alive at 03.03.2021
n = 910,262
Not eligible criteria
n = 27,120
Individuals eligible criteria
n = 890,270
Vaccinated
before 03.02.2021
n = 64,084
Individuals unvaccinated
before 03.02.2021
n = 826,186
Infected
before 03.02.2021
n = 35,407
Negative cohort
at 03.02.2021
n = 790,779
Non SNS users
n = 320,754
SNS users
n = 470,025
Unvaccinated
n = 24,003
Vaccinated
Pfizer/BioNtec
n = 379,285
Vaccinated
Moderna
n = 56,037
Vaccinated
Astrazeneca
n = 8,509
Vaccinated
Janssen
n = 2,191
